# Added value of point-of-care testing for Group A *Streptococcus* in community pharmacy sore throat pathways: Analysis of the Wales Sore Throat Test and Treat service

**DOI:** 10.64898/2026.03.18.26347584

**Authors:** Quisha Bustamante, Hannah Thornton, George Lawson, Rebecca L Guy, Haroon Ahmed, Andrew Evans, Rebecca Cannings-John, Efi Mantzourani, Christopher Jones, Colin S Brown, Victoria Hall, Theresa Lamagni, Mariyam Mirfenderesky

## Abstract

**Objective:** To evaluate the diagnostic performance of FeverPAIN and Centor with point-of-care test (POCT) results for Group A *Streptococcus* (GAS) among children and adults presenting with sore throat in community pharmacies.

**Methods:** Cross-sectional analysis of patients aged six years and over with sore throat presenting to community pharmacies across Wales delivering the Sore Throat Test and Treat (STTT) service from November 2018 to September 2024. Patients who scored FeverPAIN ≥2 or Centor ≥3 and were able to undergo POCT were eligible for analysis. We described GAS positivity by age group and assessed diagnostic performance of FeverPAIN at the National Institute for Health and Care Excellence (NICE) antibiotic threshold (≥4), reporting sensitivity, specificity, positive predictive value (PPV), negative predictive value (NPV), and area under receiver operating characteristic curve (AUROC) with 95% confidence intervals (CI). We estimated potential overtreatment and undertreatment if antibiotics were supplied based on FeverPAIN alone.

**Results:** Among 73,617 eligible patients, 37.0% (n=27,220) tested POCT-positive for GAS. Positivity was highest in children aged 6-10 years (47.0%: 5,339/11,371). FeverPAIN was used in 92.5% (n=68,099) of assessments. At the NICE-recommended threshold for antibiotic treatment (FeverPAIN ≥4), sensitivity was 55.0% (95% CI: 54.4-55.6%) and specificity 77.0% (95% CI: 76.6-77.4%). PPV was 57.6% (95% CI: 57.0-58.2%) and NPV 75.1% (95% CI: 74.7-75.5%). Overall AUROC was 0.70 (95% CI: 0.70-0.71), with the lowest AUROC of 0.69 (95% CI: 0.68-0.70) observed among children aged 6-10 years. Using FeverPAIN alone would undertreat 44% and overtreat 23% of patients based on POCT results.

**Conclusions:** FeverPAIN alone showed limited diagnostic performance for identifying GAS, with more pronounced discordance observed among children. Incorporating POCTs within community pharmacy sore throat pathways may support more targeted antibiotic prescribing. Our findings support a re-evaluation of the role of POCTs within community pharmacy sore throat pathways.

## INTRODUCTION

Acute sore throat is a common reason for primary care consultations in the UK, accounting for approximately 50 per 1,000 patients annually^1^. Although most cases are viral, antibiotics are prescribed in approximately 60% of consultations, contributing to unnecessary antimicrobial use^2^. Group A *Streptococcus* (GAS) is the main bacterial cause, but its clinical presentation overlaps with viral infections, making diagnosis challenging without confirmatory testing. Most infections are self-limiting, but can lead to local complications and invasive disease, including streptococcal toxic shock syndrome, necrotising fasciitis and bacteraemia^3^.

In England, the National Institute for Health and Care Excellence (NICE) recommends the clinical prediction rules FeverPAIN and Centor to estimate the likelihood of GAS and guide antibiotic prescribing for acute sore throat^4^. FeverPAIN, developed in 2013 in a UK primary care population, allocates one point to the presence of fever, tonsil purulence, symptom duration under three days, severely inflamed tonsils and absence of cough (maximum score of five). Centor, developed in the 1980s, scores tonsillar exudate, lymphadenopathy, fever and absence of cough (maximum score four). NICE recommends immediate or delayed antibiotics for patients scoring FeverPAIN ≥4 or Centor ≥3. However, one UK primary care study found that fewer than 25% of patients meeting these thresholds had microbiologically confirmed streptococcal infection, while some with lower scores were also positive, raising concerns about under- and overprescribing^5^.

In 2018, Wales introduced a national Sore Throat Test and Treat (STTT) service in which community pharmacists use FeverPAIN and Centor scores to determine eligibility for point-of-care testing (POCT) for GAS prior to prescribing antibiotics^6^. Northern Ireland has adopted a similar approach in its Pharmacy First service^7^. In contrast, England’s national Pharmacy First service, launched in 2024, does not include POCTs in its acute sore throat pathway for patients aged five years and over. This reflects conclusions from a 2019 NICE review which found that rapid tests for GAS in primary care were unlikely to provide additional benefit or cost-effectiveness compared with using clinical prediction rules alone^8^. The economic modelling incorporated only common suppurative complications (quinsy, otitis media, sinusitis) and did not account for invasive GAS disease or broader public health effects such as transmission and antimicrobial resistance.

The 2019 NICE review noted that diagnostic accuracy studies had not been conducted in pharmacy settings, preventing assessment of effectiveness in this setting. Since the review, studies have begun to address this evidence gap by evaluating POCT use in pharmacies. A retrospective analysis of the Wales STTT service in 2024 reported lower antibiotic supply rates than general practitioner (GP) consultations for sore throat (21% vs 39%, respectively)^9^. Additionally, a recent report of England’s Pharmacy First service reported that acute sore throat was responsible for the largest number of consultations across the scheme in 2024/25, with antimicrobial supply occurring in 65% of cases^10^. This is substantially higher than recent antibiotic supply rates reported in the Wales STTT service (29.9%) and Northern Ireland’s Pharmacy First service (25%)^7, 11^.

Given these differences in practice and emerging evidence, this study compares the diagnostic performance of FeverPAIN and Centor scores with POCT results for GAS among children and adults presenting with sore throat to the Wales STTT service.

## METHODS

### Study design

Cross-sectional study of consultations for the Wales Sore Throat Test and Treat Service.

### Participants and setting

Patients aged ≥6 years presenting with acute sore throat to the STTT service in community pharmacies across Wales between November 2018 to September 2024 were included. During this period, the service expanded to 577 community pharmacies. The protocol and data collection methods have been described previously; a summary is provided in Figure 1^6^.

**Figure 1.**
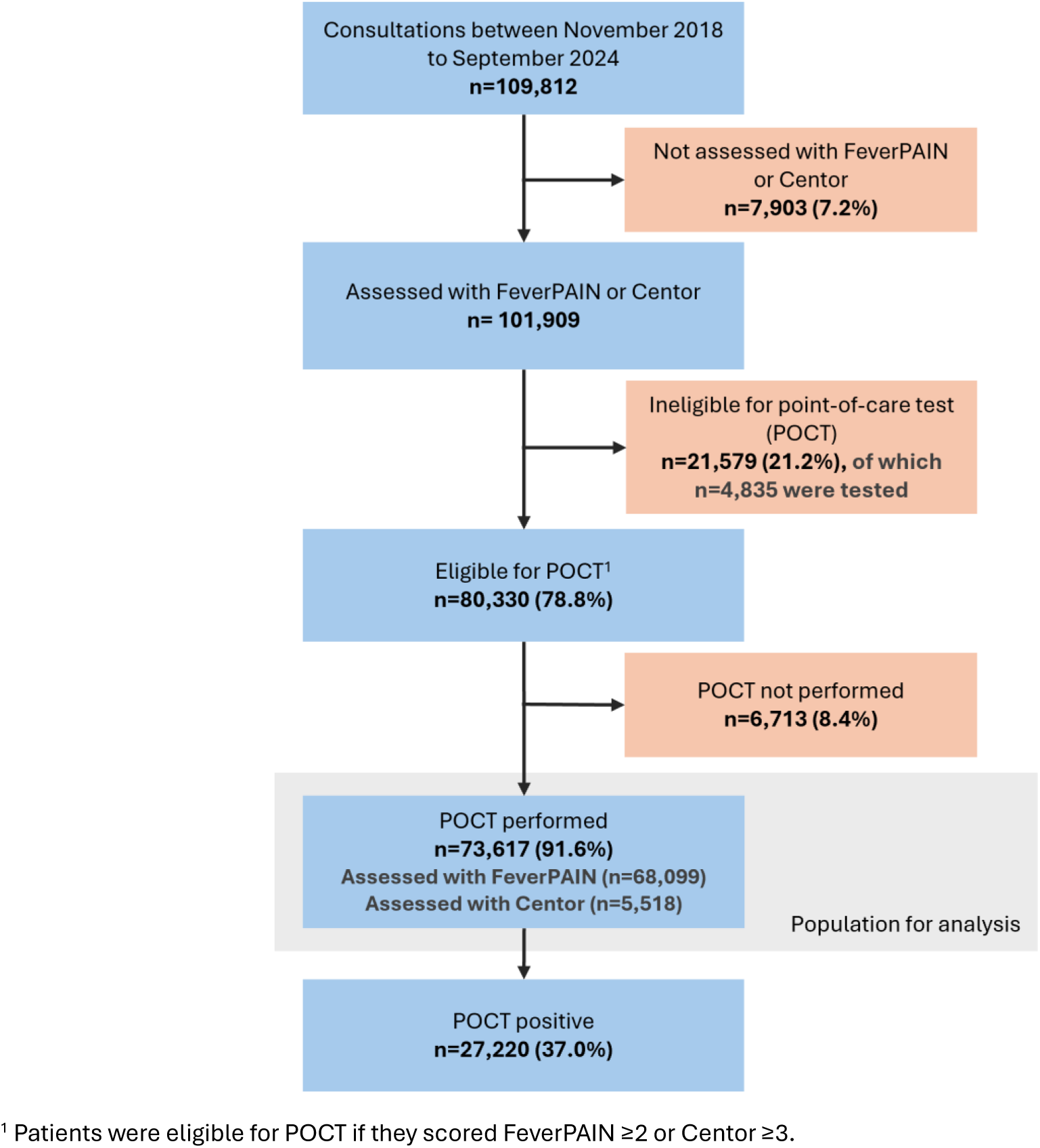
Overview of the Wales Sore Throat Test and Treat (STTT) service, November 2018 to September 2024.

Patients underwent assessment using FeverPAIN or Centor prediction rules. Patients scoring FeverPAIN ≥2 or Centor ≥3 were eligible for a POCT, administered by the pharmacist, and were supplied antibiotics if POCT-positive, following discussions with the patient. Pharmacists used the OSOM® Strep A Test (Sekisui Diagnostics), which has a reported sensitivity of 96% and specificity of 98% compared with culture^12^.

### Data sources

Consultation data were collected electronically by participating pharmacists on the Choose Pharmacy IT application. Standardised demographic information (age, sex and postcode) was obtained through linkage with the Welsh Demographic Service. Relative deprivation scores were calculated by linking patient postcodes to the Welsh Index of Multiple Deprivation (IMD) database^13^.

### Exclusion criteria

In the primary analysis, we excluded patients not assessed using FeverPAIN or Centor, those ineligible for POCT (FeverPAIN <2 or Centor <3), and those eligible for POCT but not tested.

### Primary outcomes

Key outcomes were: (1) GAS positivity by POCT and (2) diagnostic performance estimates of FeverPAIN and Centor against POCT results using sensitivity, specificity, positive predictive value (PPV), negative predictive value (NPV) and area under receiver operating characteristic (AUROC) curve.

### Primary analysis

Analyses were performed using STATA version 18.0. We summarised patient characteristics using descriptive statistics overall and by age group (6-10, 11-15, 16-40 and 41+ years). Consultation attendance and GAS positivity trends were described over time by age group.

FeverPAIN scores were analysed against the NICE antibiotic threshold (≥4). Centor could not be analysed against its NICE threshold (≥3) because POCT eligibility criteria used the same threshold, leaving no comparator group. Diagnostic performance of FeverPAIN was compared with POCT results with 95% confidence intervals (CI). GAS positivity was described across FeverPAIN scores overall and by age group.

### Sensitivity analysis

Sensitivity analyses compared characteristics of ineligible patients who underwent POCT with those who were not tested. POCT results were imputed for eligible untested patients and diagnostic performance re-estimated under two scenarios: (1) assuming the same GAS positivity as tested patients and (2) assuming all were POCT-negative. Additional sensitivity analyses included all patients who underwent POCT irrespective of eligibility criteria.

### Ethics approval

Data were collected as part of routine clinical care and fully anonymised. Data processing was undertaken under a Data Protection Impact Assessment with Digital Health and Care Wales. Ethical approval was granted by the Cardiff School of Pharmacy and Pharmaceutical Sciences Research Ethics Committee (reference number: 2324-17).

## RESULTS

### Population and service usage

A total of 109,812 STTT consultations for acute sore throat were recorded from November 2018 to September 2024. After excluding patients without a FeverPAIN or Centor assessment (7.2%: n=7,903), those ineligible for POCT (21.2%: n=21,579) and those eligible but not tested (8.4%: n=6,713), 73,617 patients were included in the analysis (Figure 1).

Mean monthly attendance was 442 consultations in 2019, increasing to 655 in 2020 before falling to six in 2021. Attendances rose in 2022 (mean of 984 consultations per month), peaking in December 2022 with 6,054 consultations, coinciding with the national surge in GAS infections, including invasive infections during the winter 2022/23 (Figure 2a)^14^. Attendance remained high thereafter as service provision expanded, with monthly means of 2,067 consultations in 2023 and 3,064 in 2024.

**Figure 2.**
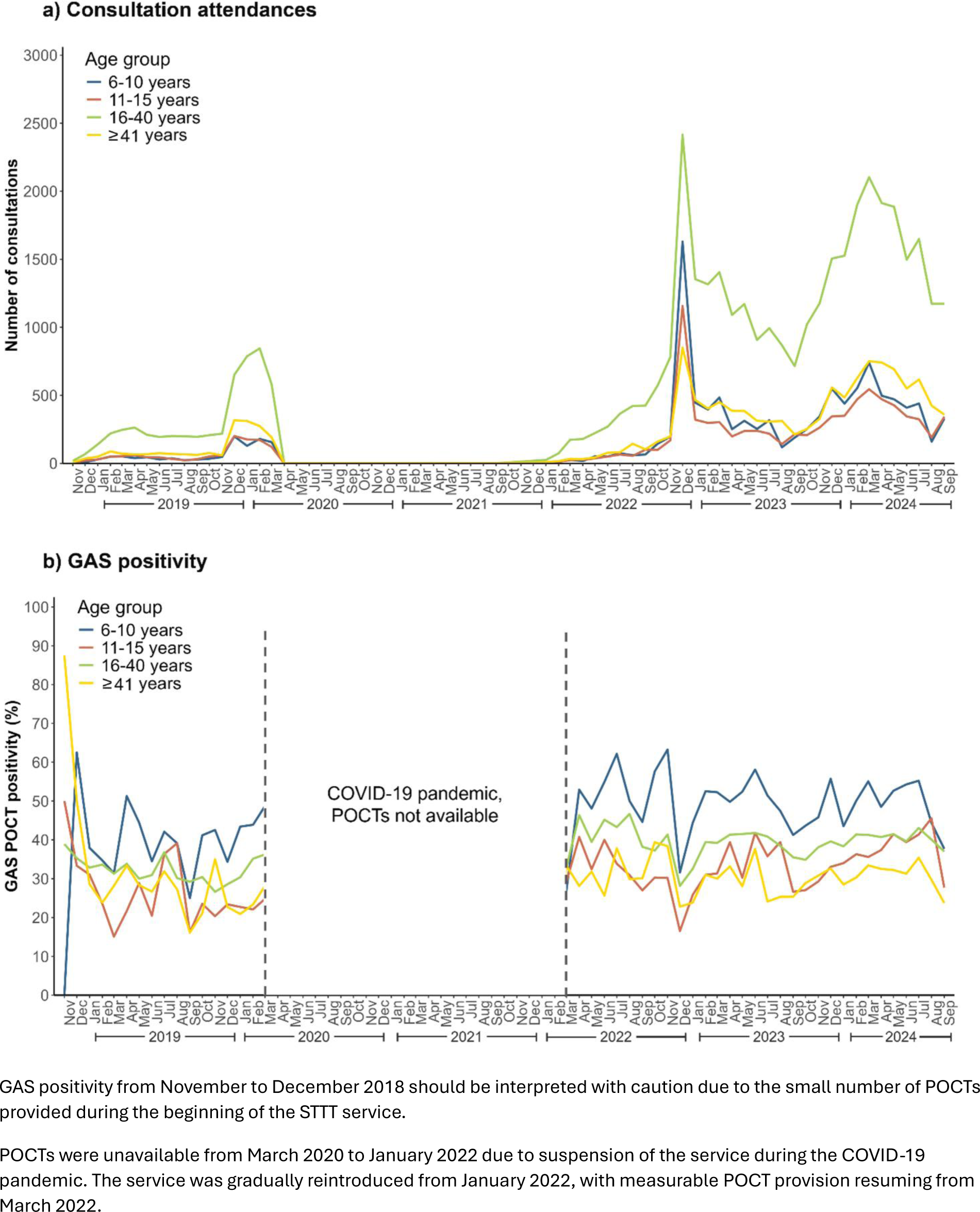
**a)** Monthly STTT consultation attendances and **b)** GAS positivity, November 2018 to September 2024.

Children under 16 years comprised 28.3% (n=20,804) of consultations and 66.7% (n=49,137) were female. A higher proportion were from the most deprived IMD quintile (21.3%) than the least deprived (15.5%) (Table 1). Self-referral was common (66.3%: n=48,785). Younger children presented earlier, with 64.5% (n=7,333/11,371) of 6-10-year-olds attending within two days of symptom onset compared with 44.1% (n=5,854/13,268) of patients aged ≥41 years. Most patients reported they would otherwise have contacted a GP (92.4%: n=67,997), while onward referral to the GP was infrequent (4.0%: n=2,949). FeverPAIN was used in 92.5% (n=68,099) of assessments and Centor in 7.5% (n=5,518).

**Table 1.**
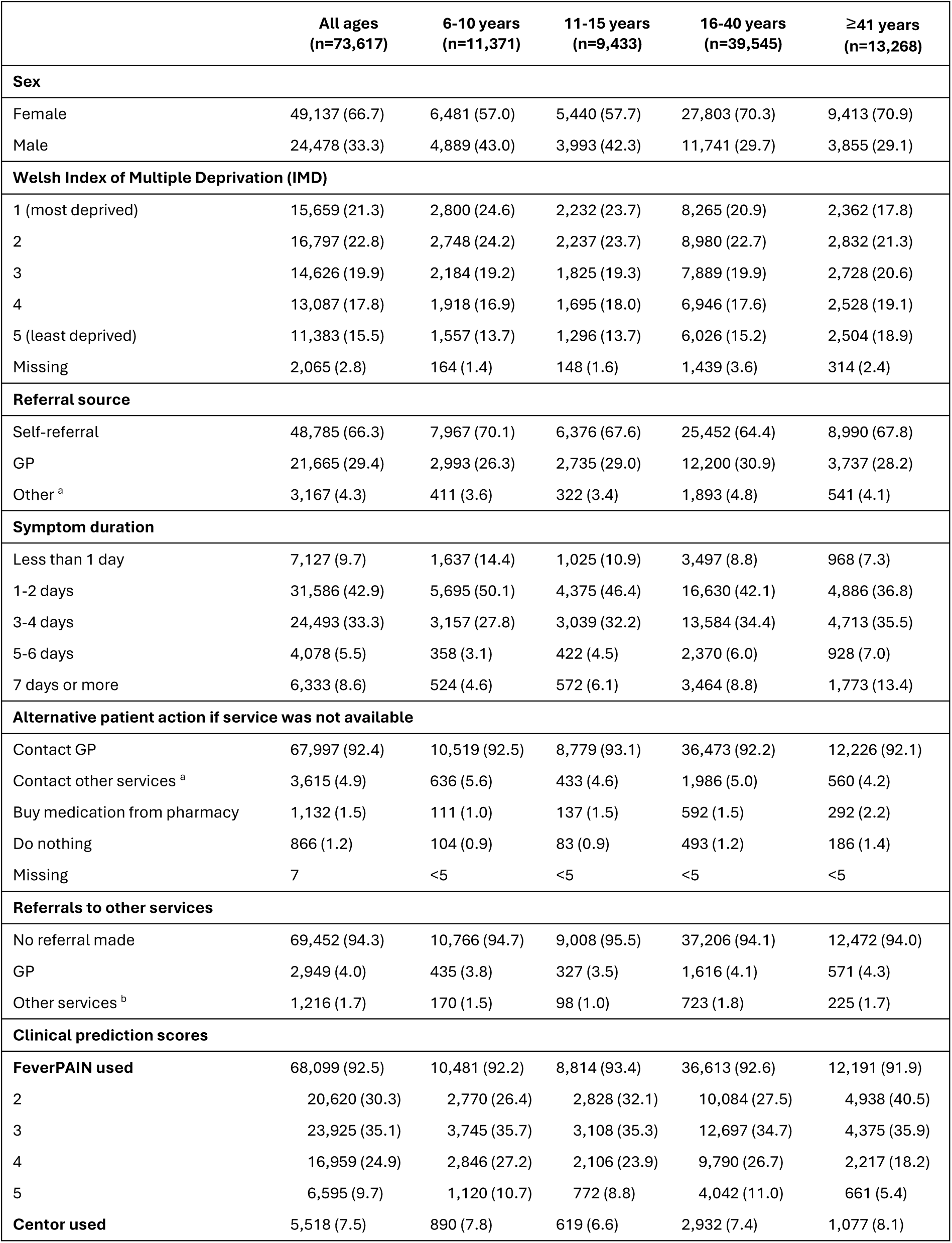
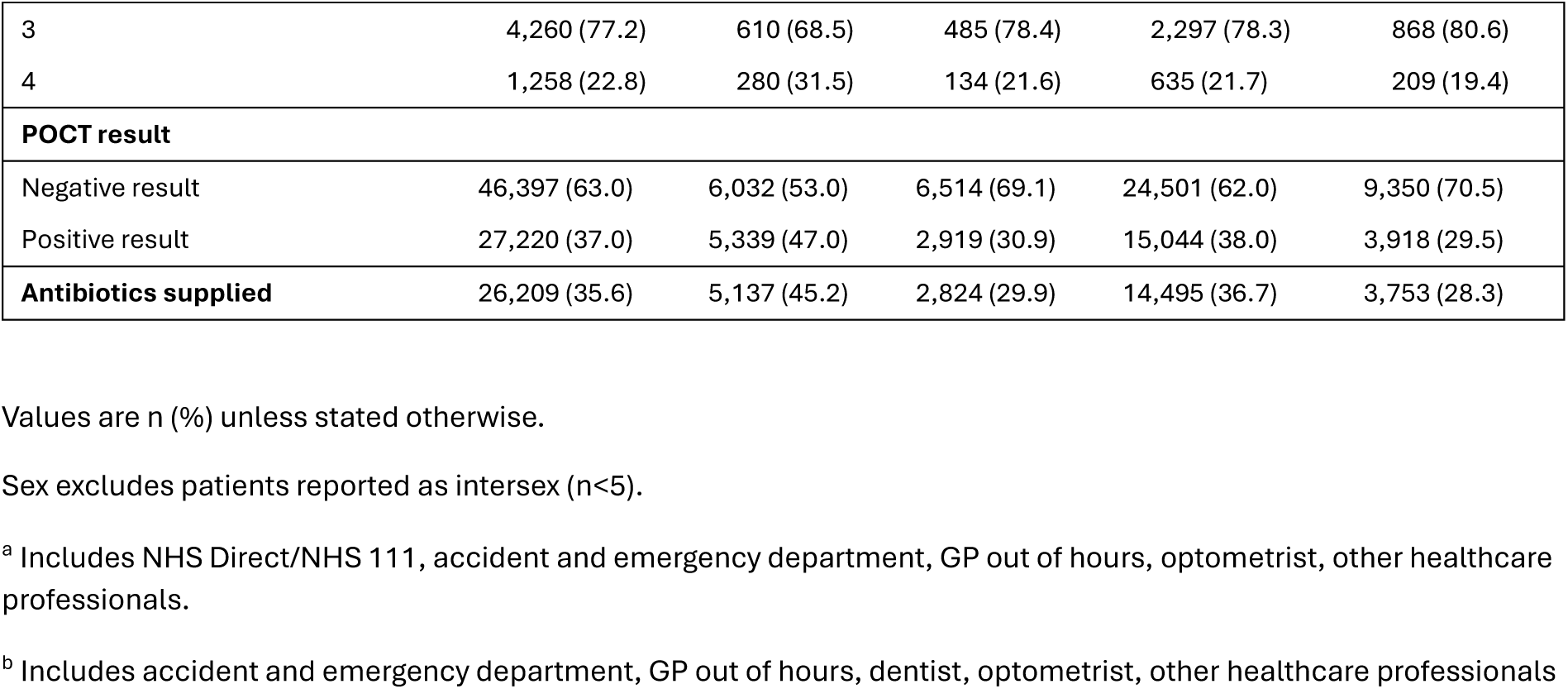
Characteristics of patients presenting to the Wales Sore Throat Test and Treat (STTT) service, November 2018 to September 2024.

|  | All ages<br>(n=73,617) | 6-10 years<br>(n=11,371) | 11-15 years<br>(n=9,433) | 16-40 years<br>(n=39,545) | ≥41 years<br>(n=13,268) |
| --- | --- | --- | --- | --- | --- |
| **Sex** |  |  |  |  |  |
| Female | 49,137 (66.7) | 6,481 (57.0) | 5,440 (57.7) | 27,803 (70.3) | 9,413 (70.9) |
| Male | 24,478 (33.3) | 4,889 (43.0) | 3,993 (42.3) | 11,741 (29.7) | 3,855 (29.1) |
| **Welsh Index of Multiple Deprivation (IMD)** |  |  |  |  |  |
| 1 (most deprived) | 15,659 (21.3) | 2,800 (24.6) | 2,232 (23.7) | 8,265 (20.9) | 2,362 (17.8) |
| 2 | 16,797 (22.8) | 2,748 (24.2) | 2,237 (23.7) | 8,980 (22.7) | 2,832 (21.3) |
| 3 | 14,626 (19.9) | 2,184 (19.2) | 1,825 (19.3) | 7,889 (19.9) | 2,728 (20.6) |
| 4 | 13,087 (17.8) | 1,918 (16.9) | 1,695 (18.0) | 6,946 (17.6) | 2,528 (19.1) |
| 5 (least deprived) | 11,383 (15.5) | 1,557 (13.7) | 1,296 (13.7) | 6,026 (15.2) | 2,504 (18.9) |
| Missing | 2,065 (2.8) | 164 (1.4) | 148 (1.6) | 1,439 (3.6) | 314 (2.4) |
| **Referral source** |  |  |  |  |  |
| Self-referral | 48,785 (66.3) | 7,967 (70.1) | 6,376 (67.6) | 25,452 (64.4) | 8,990 (67.8) |
| GP | 21,665 (29.4) | 2,993 (26.3) | 2,735 (29.0) | 12,200 (30.9) | 3,737 (28.2) |
| Other <sup>a</sup> | 3,167 (4.3) | 411 (3.6) | 322 (3.4) | 1,893 (4.8) | 541 (4.1) |
| **Symptom duration** |  |  |  |  |  |
| Less than 1 day | 7,127 (9.7) | 1,637 (14.4) | 1,025 (10.9) | 3,497 (8.8) | 968 (7.3) |
| 1-2 days | 31,586 (42.9) | 5,695 (50.1) | 4,375 (46.4) | 16,630 (42.1) | 4,886 (36.8) |
| 3-4 days | 24,493 (33.3) | 3,157 (27.8) | 3,039 (32.2) | 13,584 (34.4) | 4,713 (35.5) |
| 5-6 days | 4,078 (5.5) | 358 (3.1) | 422 (4.5) | 2,370 (6.0) | 928 (7.0) |
| 7 days or more | 6,333 (8.6) | 524 (4.6) | 572 (6.1) | 3,464 (8.8) | 1,773 (13.4) |
| **Alternative patient action if service was not available** |  |  |  |  |  |
| Contact GP | 67,997 (92.4) | 10,519 (92.5) | 8,779 (93.1) | 36,473 (92.2) | 12,226 (92.1) |
| Contact other services <sup>a</sup> | 3,615 (4.9) | 636 (5.6) | 433 (4.6) | 1,986 (5.0) | 560 (4.2) |
| Buy medication from pharmacy | 1,132 (1.5) | 111 (1.0) | 137 (1.5) | 592 (1.5) | 292 (2.2) |
| Do nothing | 866 (1.2) | 104 (0.9) | 83 (0.9) | 493 (1.2) | 186 (1.4) |
| Missing | 7 | <5 | <5 | <5 | <5 |
| **Referrals to other services** |  |  |  |  |  |
| No referral made | 69,452 (94.3) | 10,766 (94.7) | 9,008 (95.5) | 37,206 (94.1) | 12,472 (94.0) |
| GP | 2,949 (4.0) | 435 (3.8) | 327 (3.5) | 1,616 (4.1) | 571 (4.3) |
| Other services <sup>b</sup> | 1,216 (1.7) | 170 (1.5) | 98 (1.0) | 723 (1.8) | 225 (1.7) |
| **Clinical prediction scores** |  |  |  |  |  |
| **FeverPAIN used** | 68,099 (92.5) | 10,481 (92.2) | 8,814 (93.4) | 36,613 (92.6) | 12,191 (91.9) |
| 2 | 20,620 (30.3) | 2,770 (26.4) | 2,828 (32.1) | 10,084 (27.5) | 4,938 (40.5) |
| 3 | 23,925 (35.1) | 3,745 (35.7) | 3,108 (35.3) | 12,697 (34.7) | 4,375 (35.9) |
| 4 | 16,959 (24.9) | 2,846 (27.2) | 2,106 (23.9) | 9,790 (26.7) | 2,217 (18.2) |
| 5 | 6,595 (9.7) | 1,120 (10.7) | 772 (8.8) | 4,042 (11.0) | 661 (5.4) |
| **Centor used** | 5,518 (7.5) | 890 (7.8) | 619 (6.6) | 2,932 (7.4) | 1,077 (8.1) |
| 3 | 4,260 (77.2) | 610 (68.5) | 485 (78.4) | 2,297 (78.3) | 868 (80.6) |
| 4 | 1,258 (22.8) | 280 (31.5) | 134 (21.6) | 635 (21.7) | 209 (19.4) |
| **POCT result** |  |  |  |  |  |
| Negative result | 46,397 (63.0) | 6,032 (53.0) | 6,514 (69.1) | 24,501 (62.0) | 9,350 (70.5) |
| Positive result | 27,220 (37.0) | 5,339 (47.0) | 2,919 (30.9) | 15,044 (38.0) | 3,918 (29.5) |
| **Antibiotics supplied** | 26,209 (35.6) | 5,137 (45.2) | 2,824 (29.9) | 14,495 (36.7) | 3,753 (28.3) |

Values are n (%) unless stated otherwise.
Sex excludes patients reported as intersex (n<5).
<sup>a</sup> Includes NHS Direct/NHS 111, accident and emergency department, GP out of hours, optometrist, other healthcare professionals.
<sup>b</sup> Includes accident and emergency department, GP out of hours, dentist, optometrist, other healthcare professionals

### GAS positivity

Among 73,617 eligible patients, GAS positivity was 37.0% (n=27,220) with no clear temporal trend (Figure 2b). Positivity was highest among children aged 6-10 years (47.0%: n=5,339/11,371) and lowest among patients aged ≥41 years (29.5%: n=3,918/13,268). Overall, antibiotics were supplied to 35.6% (n=26,209/73,617) of patients, with the highest supply in children aged 6-10 years (45.2%: n=5,137/11,371).

### Diagnostic performance of FeverPAIN

A total of 68,099 (92.5%) patients were assessed using FeverPAIN and included in the diagnostic performance analysis; 36.2% (n=24,662) were POCT-positive. Using POCT as the service reference standard and the NICE antibiotic threshold FeverPAIN ≥4, sensitivity of FeverPAIN was 55.0% (95% CI: 54.4-55.6%) and specificity 77.0% (95% CI: 76.6-77.4%) (Table 2). The positive predictive value (PPV) was 57.6% (95% CI: 57.0-58.2%) and negative predictive value (NPV) was 75.1% (95% CI: 74.7-75.5%). Among patients scoring FeverPAIN ≥4, 57.6% were POCT-positive and 42.4% were POCT-negative; among those scoring below this threshold, 75.1% were POCT-negative and 24.9% were POCT-positive. The summary area under receiver operating characteristic (AUROC) curve was 0.70 (95% CI: 0.70-0.71) overall, with the lowest observed among children aged 6-10 years (0.69, 95% CI: 0.68-0.70) (Figure 4).

**Table 2.** Diagnostic performance of FeverPAIN score thresholds compared to POCT results (n=68,099).

| FeverPAIN threshold | Above/Below | POCT positivity (95% CI) | Sensitivity (95% CI) | Specificity (95% CI) | PPV (95% CI) | NPV (95% CI) | AUROC (95% CI) |
| --- | --- | --- | --- | --- | --- | --- | --- |
| **All ages** |  |  |  |  |  |  |  |
| ≥ 3 | 47,479/20,620 | 36.2 (35.9-36.6) | 86.0 (85.5-86.4) | 39.5 (39.0-40.0) | 44.7 (44.2-45.1) | 83.2 (82.7-83.7) | 0.63 (0.62-0.63) |
| ≥ 4 | 23,554/44,545 |  | 55.0 (54.4-55.6) | 77.0 (76.6-77.4) | 57.6 (57.0-58.2) | 75.1 (74.7-75.5) | 0.66 (0.66-0.66) |
| 5 | 6,595/61,504 |  | 18.9 (18.4-19.4) | 95.6 (95.4-95.7) | 70.7 (69.6-71.8) | 67.5 (67.1-67.9) | 0.57 (0.57-0.57) |
| **6-10 years** |  |  |  |  |  |  |  |
| ≥ 3 | 7,711/2,770 | 45.8 (44.8-46.7) | 85.5 (84.5-86.5) | 36.5 (35.2-37.8) | 53.2 (52.1-54.3) | 74.9 (73.2-76.5) | 0.61 (0.60-0.62) |
| ≥ 4 | 3,966/6,515 |  | 54.2 (52.8-55.6) | 76.0 (74.9-77.1) | 65.6 (64.1-67.1) | 66.3 (65.1-67.4) | 0.65 (0.64-0.66) |
| 5 | 1,120/9,361 |  | 18.3 (17.2-19.4) | 95.7 (95.1-96.2) | 78.2 (75.7-80.6) | 58.1 (57.1-59.1) | 0.57 (0.56-0.58) |
| **11-15 years** |  |  |  |  |  |  |  |
| ≥ 3 | 5,986/2,828 | 30.3 (29.4-31.3) | 87.0 (85.6-88.2) | 40.4 (39.1-41.6) | 38.8 (37.6-40.1) | 87.7 (86.4-88.9) | 0.64 (0.63-0.65) |
| ≥ 4 | 2,878/5,936 |  | 56.9 (55.0-58.8) | 77.9 (76.8-78.9) | 52.8 (51.0-54.7) | 80.6 (79.6-81.6) | 0.67 (0.66-0.68) |
| 5 | 772/8,042 |  | 20.3 (18.8-21.9) | 96.3 (95.7-96.7) | 70.2 (66.8-73.4) | 73.5 (72.5-74.5) | 0.58 (0.57-0.59) |
| **16-40 years** |  |  |  |  |  |  |  |
| ≥ 3 | 26,529/10,084 | 37.4 (36.9-37.9) | 87.3 (86.8-87.9) | 36.4 (35.8-37.0) | 45.0 (44.4-45.6) | 82.8 (82.1-83.5) | 0.62 (0.61-0.62) |
| ≥ 4 | 13,832/22,781 |  | 57.3 (56.5-58.1) | 73.9 (73.3-74.4) | 56.7 (55.9-57.5) | 74.4 (73.8-74.9) | 0.66 (0.65-0.66) |
| 5 | 4,042/32,571 |  | 20.2 (19.6-20.9) | 94.5 (94.1-94.7) | 68.5 (67.1-70.0) | 66.5 (66.0-67.0) | 0.57 (0.57-0.58) |
| **≥41 years** |  |  |  |  |  |  |  |
| ≥ 3 | 7,253/4,938 | 28.8 (28.0-29.6) | 80.6 (79.3-81.9) | 49.0 (48.0-50.1) | 39.0 (37.9-40.1) | 86.2 (85.2-87.2) | 0.65 (0.64-0.66) |
| ≥ 4 | 2,878/9,313 |  | 45.7 (44.0-47.3) | 85.3 (84.5-86.0) | 55.7 (53.8-57.5) | 79.5 (78.7-80.3) | 0.65 (0.65-0.66) |
| 5 | 661/11,530 |  | 13.6 (12.4-14.7) | 97.9 (97.5-98.2) | 72.0 (68.4-75.4) | 73.7 (72.9-74.5) | 0.56 (0.55-0.56) |

Above/Below = consultations above FeverPAIN threshold/consultations below FeverPAIN threshold. PPV = positive predictive value. NPV = negative predictive value. AUROC = area under receiver operating characteristic curve.

### Association between FeverPAIN score and GAS positivity

Figure 3 shows a positive association between FeverPAIN score and GAS positivity across all age groups. POCT-positivity increased from 16.8% at FeverPAIN 2 to 70.7% at FeverPAIN 5. Children aged 6-10 years consistently had the highest positivity at each FeverPAIN score.

**Figure 3.**
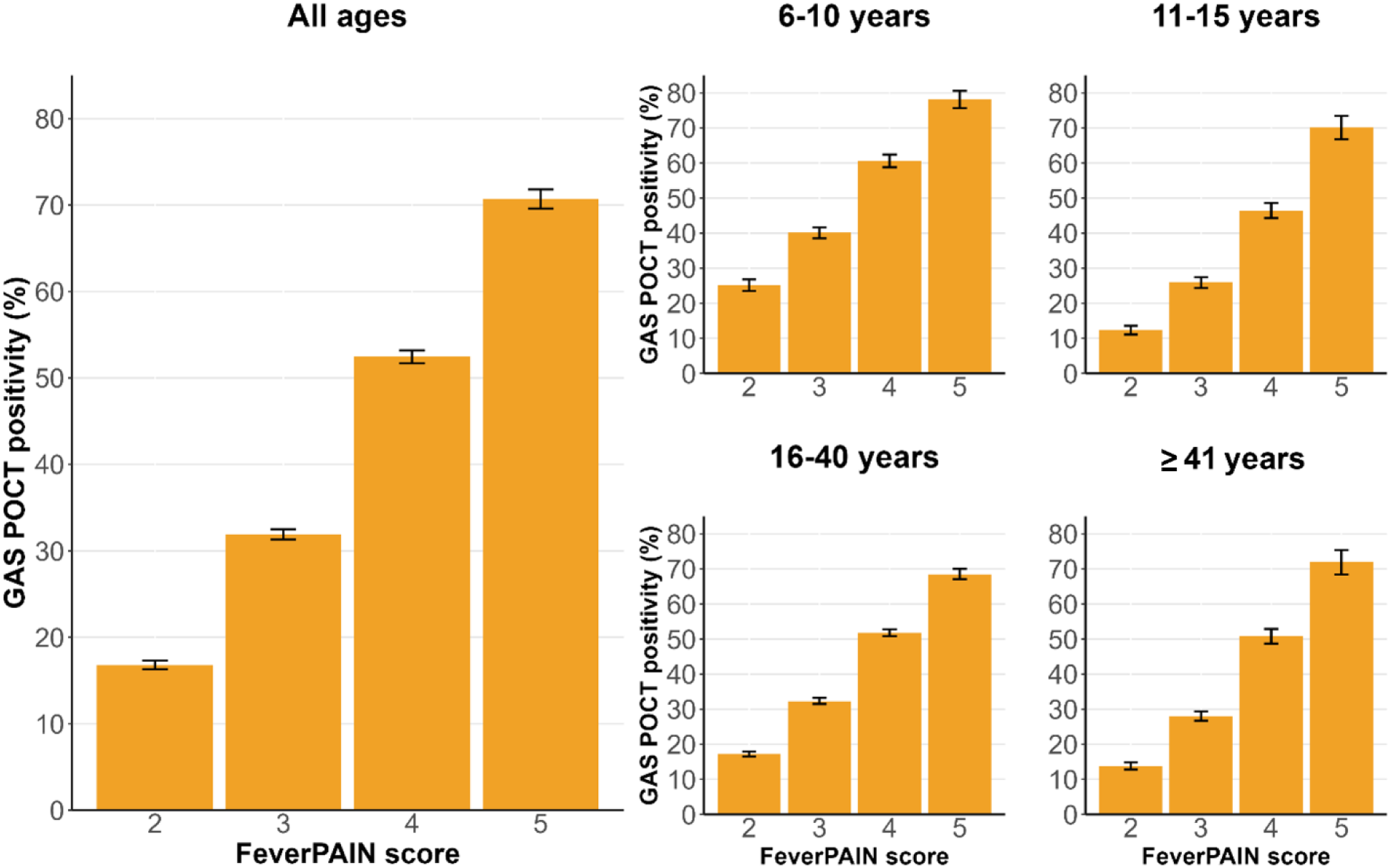
GAS positivity rate and 95% confidence intervals for FeverPAIN scores by age group.

**Figure 4.**
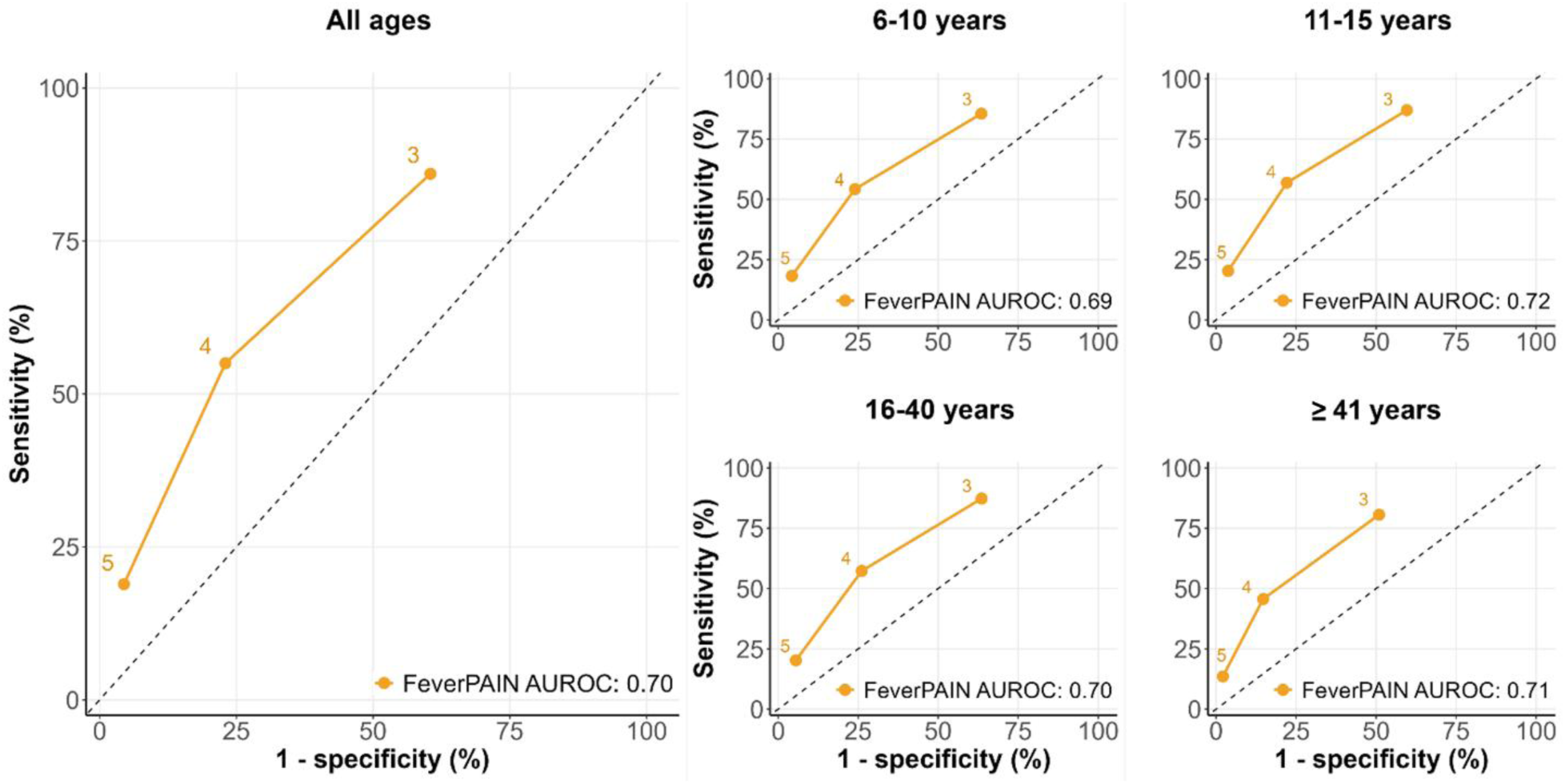
Receiver operating characteristic curves for FeverPAIN thresholds by age group.

### Clinical implications of using FeverPAIN alone

Based on an observed GAS positivity of 36.2%, approximately 36 of every 100 patients presenting to the pharmacy service would be POCT-positive and 64 POCT-negative patients. If antibiotic supply were based solely on NICE guidance (FeverPAIN ≥4), an estimated 16 of the 36 POCT-positive patients (44%) would not receive antibiotics, while approximately 15 of the 64 POCT-negative patients (23%) would receive antibiotics despite no evidence of GAS infection (Figure 5). By age group, missed antibiotic treatment was most pronounced among children aged 6-10 years, where 21 of 46 POCT-positive patients (46%) would not receive antibiotics (Supplementary Figure 1). Potential overtreatment was highest in patients aged 16-40 years, where 16 of 63 POCT-negative patients (25%) would receive antibiotics.

**Figure 5.**
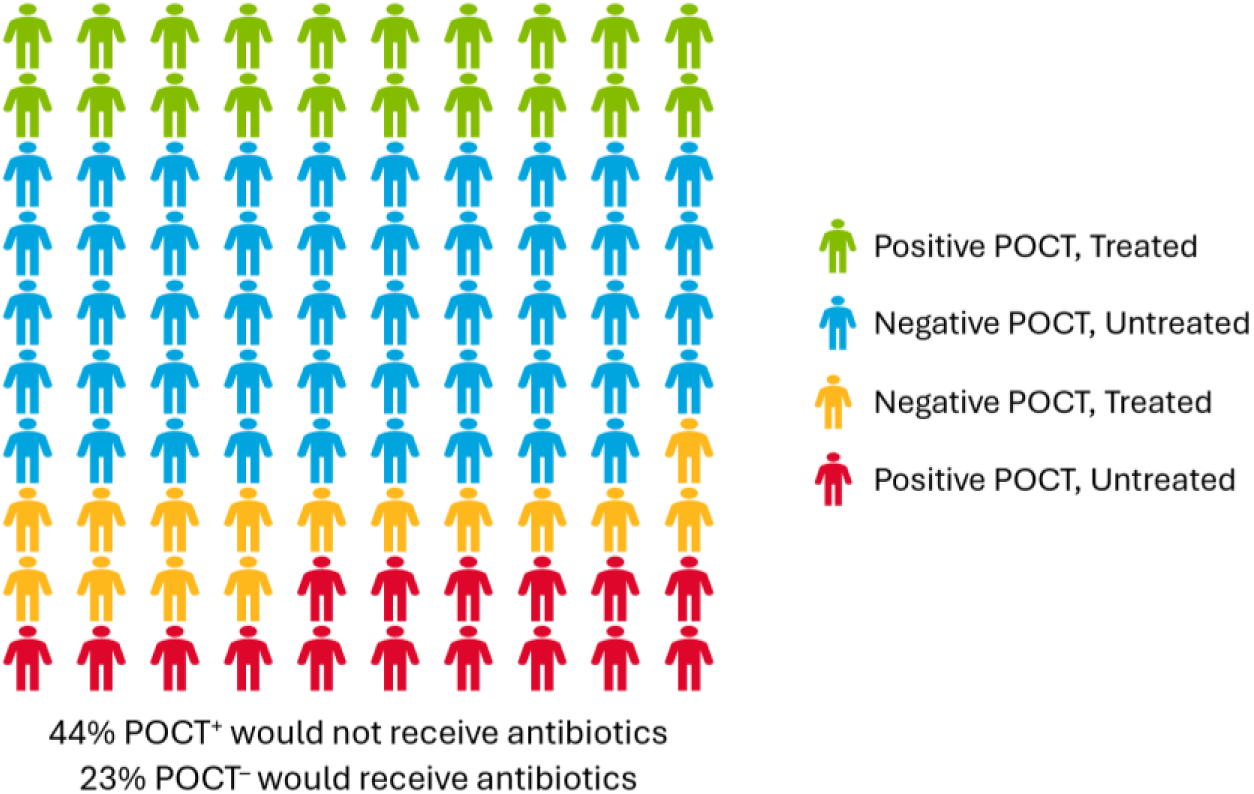
Proportion of treated and untreated patients if FeverPAIN ≥4 alone was used to guide antibiotic prescribing in 100 patients presenting with sore throat to the STTT service using a GAS positivity of 36%.

### Sensitivity analysis

A total of 4,835 patients ineligible for POCT were tested; reasons for testing were unavailable. Among these, 12.3% (n=593) were POCT-positive for GAS (Supplementary Table 1).

After imputing POCT results for 6,462 eligible patients who did not undergo testing, the analytic sample increased from 68,099 to 74,561, with no material change in the diagnostic performance estimates (Supplementary Table 2a and 2b). Analyses including all patients who underwent a POCT, including those below the eligibility criteria (n=4,835), produced comparable estimates (Supplementary Table 3).

## DISCUSSION

Analysis of data from the Wales STTT service indicates that clinical prediction rules alone cannot reliably distinguish GAS infection in community pharmacy sore throat pathways. Among patients meeting the NICE threshold for antibiotic prescribing (FeverPAIN ≥4), 42% tested POCT-negative, while 25% below the threshold were POCT-positive, indicating potential overtreatment and missed treatment if FeverPAIN were used in isolation.

POCT positivity increased with higher FeverPAIN scores, broadly consistent with NICE (NG84) estimates derived largely from throat culture-based studies, where scores of 4–5 are associated with a 62–65% probability of streptococcal isolation and scores of 0–1 with probabilities below 20%^5^. In this cohort, POCT positivity increased from 16.8% at a score of 2 to 70.7% at a score of 5. However, overlap between adjacent scores was greater than those suggested by guideline estimates, indicating that while FeverPAIN may be calibrated at a population level, it lacks sufficient precision for individual decision-making without confirmatory testing.

Younger children demonstrated the highest GAS positivity across all FeverPAIN scores but also the greatest discordance between clinical score and POCT result. Conversely, adults were more likely to be POCT-negative despite meeting the treatment threshold, increasing the risk of unnecessary antibiotic prescribing. Taken together, these findings highlight limitations in applying GP-derived prediction rules within high-throughput community pharmacy sore throat pathways, where protocol-driven assessments may provide limited scope for clinical judgement, particularly during periods of increased GAS incidence when pre-test probability is higher. In this context, incorporating POCTs within pharmacy pathways may improve diagnostic accuracy and support more targeted antibiotic use across age groups.

Previous evaluations of the STTT service have highlighted limitations of relying solely on clinical prediction rules. Mantzourani *et al*. reported that over one-third of patients meeting NICE thresholds for antibiotics were POCT-negative, while more than a quarter below the threshold were POCT-positive, illustrating the risk of misclassification if clinical prediction rules were used in isolation^11^. Our findings align with and extend these observations by quantifying the diagnostic performance and its implications for antibiotic prescribing across age groups.

Reported antibiotic prescribing rates for sore throat consultations vary across UK community pharmacy models (Wales: 29.9%, Northern Ireland: 25%, England: 65.5%)^7, 10, 11^. While comparisons should be interpreted cautiously given differences in pathway design and patient populations, this variation may reflect the availability of confirmatory POCT. This is supported by observations from Wales during the temporary suspension of GAS POCT in the COVID-19 pandemic (March 2020 to January 2022), when pharmacist prescribing increased from 27% to 63%, despite national surveillance data showing exceptionally low levels of GAS circulation compared with the pre-pandemic average^15, 16^.

The principal strength of this study is the large sample size derived from a real-world national dataset spanning 577 community pharmacies across Wales, supporting the external validity of the findings. The use of routinely collected clinical data enhances the applicability to clinical practice. Age-stratified analyses enabled assessment of diagnostic performance across age groups, including children, in whom GAS infection is most common. Minimal missing data and consistent sensitivity analyses further strengthen the robustness of our findings.

This study has several limitations. The absence of a microbiological reference standard, such as throat culture, limits certainty regarding true infection status. POCTs were used as a pragmatic proxy for microbiological confirmation within routine practice. Although not a definitive gold standard, validated GAS POCT assays have established diagnostic performance and provide timely information to support antibiotic decision-making in community pharmacies^12^.

This study did not distinguish between active infection and asymptomatic carriage. GAS carriage is recognised, although prevalence varies across age-groups and settings. A 2018 meta-analysis estimated paediatric carriage at 10.5%, whereas more recent London data reported a lower non-outbreak carriage rate of 2.8%^17, 18^.

As participants were symptomatic at presentation, detection of GAS by POCT may be more likely to represent clinical infection rather than incidental carriage, although this cannot be confirmed with the available dataset. In routine practice, GAS detection in a patient presenting with sore throat would not typically be disregarded as carriage alone. Notably, POCT availability in this analysis was associated with lower, rather than higher, antibiotic prescribing, which does not support widespread treatment of incidental carriage. Alternative pathogens were not systematically tested for and clinical outcomes, including complications, onward referral, and healthcare utilisation, were not assessed.

Measures of FeverPAIN performance in this study should be interpreted within the higher-risk population eligible for POCT rather than the full spectrum of sore throat presentations. Even within this group, discordance between FeverPAIN score and POCT result remained evident.

Future research should include economic evaluation of POCT integration into community pharmacy sore throat pathways, including the consequences of misclassification such as unnecessary antibiotic exposure and missed or delayed treatment. Evaluations should adopt a health-system perspective and assess downstream outcomes including repeat consultations, escalation of care, and antibiotic-related adverse events. Given the greater discordance observed in children, further evaluation in paediatric populations and assessment of wider public health impacts, including antimicrobial resistance, antibiotic consumption and GAS transmission dynamics are also warranted.

In conclusion, within community pharmacy sore throat pathways, FeverPAIN demonstrated limited diagnostic performance for identifying GAS when used as a standalone tool, with greater discordance observed in younger children. Integration of POCTs within community pharmacy services may improve identification of GAS infection and support more targeted antibiotic prescribing. These findings support reconsideration of the role of POCTs in acute sore throat management within community pharmacy settings, with a view to wider adoption.

## TRANSPARENCY DECLARATION

### Access to data

Anonymised data can be shared upon reasonable request and in accordance with organisational data sharing governance arrangements.

### CRediT authorship contribution statement

QB: Formal Analysis, Visualisation, Writing – Original Draft, Writing – Review & Editing. HT: Conceptualisation, Methodology, Writing – Original Draft, Writing – Review & Editing. GL: Writing – Review & Editing. RLG: Writing Review & Editing. HA: Writing – Review & Editing. AE: Writing – Review & Editing. RC: Formal Analysis, Writing Review & Editing. EM: Writing – Review & Editing, Supervision. CJ: Writing – Review & Editing. CSB: Supervision, Writing – Review & Editing. VH: Conceptualisation, Writing – Review & Editing, Supervision. TL: Conceptualisation, Writing – Review & Editing, Supervision. MM: Conceptualisation, Writing – Review & Editing, Supervision.

### Declaration of generative AI use

Generative AI tools were used to assist with structural refinement and language editing. All scientific interpretation, analysis and references were developed and verified by the author team.

## Acknowledgements

We would like to thank the Support and Business Analysts in Digital Health and Care Wales for their support with obtaining the STTT data.

