## Supplementary Tables and Figures for "Added value of point-of-care testing for Group A *Streptococcus* in community pharmacy sore throat pathways: Analysis of the Wales Sore Throat Test and Treat service"

**Supplementary Table 1.** Characteristics of POCT ineligible patients who were tested by POCT (n=4,835) versus not tested (n=16,744).

| | Ineligible and tested (n=4,835) | Ineligible and untested (n=16,744) |
|---|---|---|
| **Age group** | | |
| 6-10 years | 577 (11.9) | 2,179 (13.0) |
| 11-15 years | 595 (12.3) | 2,188 (13.1) |
| 16-40 years | 2,162 (44.7) | 6,632 (39.6) |
| ≥41 years | 1,501 (31.0) | 5,745 (34.3) |
| **Sex** | | |
| Female | 3,276 (67.8) | 10,888 (65.0) |
| Male | 1,559 (32.2) | 5,854 (35.0) |
| **Index of Multiple Deprivation (IMD)** | | |
| 1 (most deprived) | 916 (18.9) | 3,892 (23.2) |
| 2 | 1,187 (24.6) | 3,894 (23.3) |
| 3 | 1,138 (23.5) | 3,376 (20.2) |
| 4 | 822 (17.0) | 2,913 (17.4) |
| 5 (least deprived) | 652 (13.5) | 2,332 (13.9) |
| Missing | 120 (2.5) | 337 (2.0) |
| **Referral source** | | |
| Self-referral | 3,686 (76.2) | 11,570 (69.1) |
| GP | 997 (20.6) | 4,637 (27.7) |
| Other [a] | 152 (3.1) | 537 (3.2) |
| **Symptom duration** | | |
| Less than 1 day | 178 (3.7) | 736 (4.4) |
| 1-2 days | 797 (16.5) | 2,838 (16.9) |
| 3-4 days | 1,490 (30.8) | 4,983 (29.8) |
| 5-6 days | 771 (15.9) | 2,370 (14.2) |
| 7 days or more | 1,599 (33.1) | 5,817 (34.7) |
| **Alternative patient action if service was not available** | | |
| Contact GP | 4,371 (90.4) | 14,971 (89.4) |
| Contact other services [a] | 153 (3.2) | 573 (3.4) |
| Bought medication from pharmacy | 183 (3.8) | 817 (4.9) |
| Do nothing | 128 (2.6) | 381 (2.3) |
| Missing | - | <5 |
| **Referrals to other services** | | |
| No referral made | 4,471 (92.5) | 14,355 (85.7) |
| GP | 285 (5.9) | 1,895 (11.3) |
| Other services [b] | 79 (1.6) | 494 (3.0) |
| **Clinical prediction scores** | | |
| **FeverPAIN used** | 3,654 (75.6) | 13,902 (83.0) |
| 0 | 724 (19.8) | 5,227 (37.6) |
| 1 | 2,928 (80.1) | 8,675 (62.4) |
| Missing | <5 | - |
| **Centor used** | 1,181 (24.4) | 2,842 (17.0) |
| 0 | 52 (4.4) | 639 (22.5) |
| 1 | 272 (23.0) | 1,313 (46.2) |
| 2 | 857 (72.6) | 890 (31.3) |
| **POCT result** | | |
| Negative result | 4,242 (87.7) | - |
| Positive result | 593 (12.3) | - |
| **Antibiotics supplied** | 547 (11.3) | 12 (0.1) |

Values are n (%) unless stated otherwise.

Sex excludes patients reported as intersex (n<5).

[a] Includes NHS Direct/NHS 111, accident and emergency department, GP out of hours, optometrist, other healthcare professionals.

[b] Includes accident and emergency department, out of hours, dentist, optometrist, other healthcare professionals.

**Supplementary Table 2a.** Diagnostic performance of FeverPAIN score thresholds compared to POCT result under the assumption that untested patients (n=6,462) had the same proportion of POCT-positive as those tested (total n=74,561).

| FeverPAIN threshold | Above/Below | POCT positivity (95% CI) | Sensitivity (95% CI) | Specificity (95% CI) | PPV (95% CI) | NPV (95% CI) | AUROC (95% CI) |
|---|---|---|---|---|---|---|---|
| **All ages** | | | | | | | |
| **≥ 3** | 49,960/24,601 | 35.4 (35.1-35.8) | 84.4 (83.9-84.8) | 42.5 (42.1-43.0) | 44.6 (44.2-45.1) | 83.2 (82.8-83.7) | 0.63 (0.63-0.64) |
| **≥ 4** | 24,662/49,899 | | 53.8 (53.2-54.4) | 78.3 (77.9-78.7) | 57.6 (57.0-58.3) | 75.5 (75.2-75.9) | 0.66 (0.66-0.66) |
| **5** | 6,977/67,584 | | 18.7 (18.2-19.2) | 95.8 (95.6-95.9) | 70.7 (69.6-71.8) | 68.2 (67.9-68.6) | 0.57 (0.57-0.57) |
| **6-10 years** | | | | | | | |
| **≥ 3** | 8,306/3,538 | 43.7 (42.8-44.5) | 84.0 (83.0-85.0) | 40.6 (39.5-41.8) | 52.3 (51.2-53.4) | 76.7 (75.2-78.0) | 0.62 (0.62-0.63) |
| **≥ 4** | 4,215/7,629 | | 52.8 (51.5-54.2) | 77.8 (76.8-78.8) | 64.8 (63.4-66.3) | 68.0 (67.0-69.1) | 0.65 (0.64-0.66) |
| **5** | 1,196/10,648 | | 17.9 (16.9-19.0) | 95.9 (95.4-96.4) | 77.3 (74.9-79.7) | 60.1 (59.2-61.1) | 0.57 (0.56-0.57) |
| **11-15 years** | | | | | | | |
| **≥ 3** | 6,298/3,429 | 30.0 (29.1-30.9) | 84.6 (83.2-85.9) | 43.7 (42.5-44.9) | 39.1 (37.9-40.4) | 86.9 (85.7-88.0) | 0.64 (0.63-0.65) |
| **≥ 4** | 2,995/6,732 | | 54.8 (53.0-56.6) | 79.5 (78.5-80.4) | 53.3 (51.5-55.1) | 80.4 (79.5-81.4) | 0.67 (0.66-0.68) |
| **5** | 820/8,907 | | 19.8 (18.4-21.3) | 96.4 (96.0-96.9) | 70.4 (67.1-73.5) | 73.8 (72.8-74.7) | 0.58 (0.57-0.59) |
| **16-40 years** | | | | | | | |
| **≥ 3** | 27,707/11,709 | 36.7 (36.2-37.2) | 86.2 (85.6-86.8) | 38.9 (38.3-39.5) | 45.0 (44.4-45.6) | 82.9 (82.3-83.6) | 0.63 (0.62-0.63) |
| **≥ 4** | 14,435/24,981 | | 56.6 (55.8-57.4) | 75.0 (74.4-75.5) | 56.7 (55.9-57.6) | 74.8 (74.3-75.4) | 0.66 (0.65-0.66) |
| **5** | 4,256/35,160 | | 20.2 (19.5-20.9) | 94.7 (94.4-94.9) | 68.7 (67.2-70.0) | 67.1 (66.7-67.6) | 0.57 (0.57-0.58) |
| **≥41 years** | | | | | | | |
| **≥ 3** | 7,649/5,925 | 28.4 (27.7-29.2) | 77.9 (76.5-79.2) | 52.2 (51.2-53.2) | 39.3 (38.2-40.4) | 85.6 (84.7-86.5) | 0.65 (0.64-0.66) |
| **≥ 4** | 3,017/10,557 | | 44.0 (42.4-45.5) | 86.4 (85.7-87.1) | 56.2 (54.5-58.0) | 79.5 (78.7-80.3) | 0.65 (0.64-0.66) |
| **5** | 705/12,869 | | 13.2 (12.2-14.3) | 98.0 (97.7-98.3) | 72.3 (68.9-75.6) | 74.0 (73.2-74.7) | 0.56 (0.55-0.56) |

Above/Below = consultations above FeverPAIN threshold/consultations below FeverPAIN threshold. PPV = positive predictive value. NPV = negative predictive value. AUROC = area under receiver operating characteristic curve.

**Supplementary Table 2b.** Diagnostic performance of FeverPAIN score thresholds compared to POCT results under the assumption that untested patients (n=6,462) were POCT-negative (total n=74,561).

| FeverPAIN threshold | Above/Below | POCT positivity (95% CI) | Sensitivity (95% CI) | Specificity (95% CI) | PPV (95% CI) | NPV (95% CI) | AUROC (95% CI) |
|---|---|---|---|---|---|---|---|
| **All ages** | | | | | | | |
| **≥ 3** | 49,960/24,601 | 33.1 (32.7-33.4) | 86.0 (85.5-86.4) | 42.4 (41.9-42.8) | 42.4 (42.0-42.9) | 85.9 (85.5-86.4) | 0.64 (0.64-0.64) |
| **≥ 4** | 24,662/49,899 | | 55.0 (54.4-55.6) | 77.8 (77.4-78.1) | 55.0 (54.4-55.6) | 77.8 (77.4-78.1) | 0.66 (0.66-0.67) |
| **5** | 6,977/67,584 | | 18.9 (18.4-19.4) | 95.4 (95.2-95.5) | 66.8 (65.7-68.0) | 70.4 (70.1-70.8) | 0.57 (0.57-0.57) |
| **6-10 years** | | | | | | | |
| **≥ 3** | 8,306/3,538 | 40.5 (39.6-41.4) | 85.5 (84.5-86.5) | 40.3 (39.2-41.5) | 49.4 (48.3-50.5) | 80.3 (79.0-81.6) | 0.63 (0.62-0.64) |
| **≥ 4** | 4,215/7,629 | | 54.2 (52.8-55.6) | 77.1 (76.1-78.1) | 61.7 (60.2-63.2) | 71.2 (70.2-72.2) | 0.66 (0.65-0.67) |
| **5** | 1,196/10,648 | | 18.3 (17.2-19.4) | 95.5 (94.9-95.9) | 73.2 (70.6-75.7) | 63.2 (62.2-64.1) | 0.57 (0.56-0.57) |
| **11-15 years** | | | | | | | |
| **≥ 3** | 6,298/3,429 | 27.5 (26.6-28.4) | 87.0 (85.6-88.2) | 43.7 (42.5-44.8) | 36.9 (35.7-38.1) | 89.9 (88.8-90.8) | 0.65 (0.64-0.66) |
| **≥ 4** | 2,995/6,732 | | 56.9 (55.0-58.8) | 79.1 (78.1-80.0) | 50.8 (48.9-52.6) | 82.9 (82.0-83.8) | 0.68 (0.67-0.69) |
| **5** | 820/8,907 | | 20.3 (18.8-21.9) | 96.1 (95.6-96.5) | 66.1 (62.7-69.3) | 76.1 (75.2-77.0) | 0.58 (0.57-0.59) |
| **16-40 years** | | | | | | | |
| **≥ 3** | 27,707/11,709 | 34.7 (34.2-35.2) | 87.3 (86.8-87.9) | 38.8 (38.2-39.4) | 43.1 (42.5-43.7) | 85.2 (84.5-85.8) | 0.63 (0.63-0.63) |
| **≥ 4** | 14,435/24,981 | | 57.3 (56.5-58.1) | 74.4 (73.8-74.9) | 54.3 (53.5-55.1) | 76.6 (76.1-77.1) | 0.66 (0.65-0.66) |
| **5** | 4,256/35,160 | | 20.2 (19.6-20.9) | 94.2 (93.9-94.5) | 65.1 (63.6-66.5) | 69.0 (68.5-69.4) | 0.57 (0.57-0.58) |
| **≥41 years** | | | | | | | |
| **≥ 3** | 7,649/5,925 | 25.9 (25.1-26.6) | 80.6 (79.3-81.9) | 52.1 (51.1-53.1) | 37.0 (35.9-38.1) | 88.5 (87.7-89.3) | 0.66 (0.66-0.67) |
| **≥ 4** | 3,017/10,557 | | 45.7 (44.0-47.3) | 85.9 (85.2-86.6) | 53.1 (51.3-54.9) | 81.9 (81.2-82.7) | 0.66 (0.65-0.67) |
| **5** | 705/12,869 | | 13.6 (12.4-14.7) | 97.7 (97.4-98.0) | 67.5 (63.9-71.0) | 76.4 (75.7-77.2) | 0.56 (0.55-0.56) |

Above/Below = consultations above FeverPAIN threshold/consultations below FeverPAIN threshold. PPV = positive predictive value. NPV = negative predictive value. AUROC = area under receiver operating characteristic curve.

**Supplementary Table 3.** Diagnostic performance of FeverPAIN (n=71,751) and Centor (n=6,699) score thresholds compared to POCT results if all tested patients were used in analysis (total n=78,450).

| Threshold | Above/Below | POCT positivity (95% CI) | Sensitivity (95% CI) | Specificity (95% CI) | PPV (95% CI) | NPV (95% CI) | AUROC (95% CI) |
|---|---|---|---|---|---|---|---|
| **All Ages** | | | | | | | |
| **FeverPAIN** | | | | | | | |
| **≥ 1** | 71,027/724 | 34.9 (34.5-35.2) | 99.8 (99.8-99.9) | 1.5 (1.4-1.6) | 35.2 (34.8-35.5) | 94.8 (92.9-96.3) | 0.51 (0.51-0.51) |
| **≥ 2** | 68,099/3,652 | | 98.6 (98.4-98.7) | 7.0 (6.8-7.3) | 36.2 (35.9-36.6) | 90.1 (89.1-91.1) | 0.53 (0.53-0.53) |
| **≥ 3** | 47,479/24,272 | | 84.7 (84.3-85.2) | 43.8 (43.3-44.2) | 44.7 (44.2-45.1) | 84.3 (83.8-84.7) | 0.64 (0.64-0.65) |
| **≥ 4** | 23,554/48,197 | | 54.2 (53.6-54.8) | 78.6 (78.3-79.0) | 57.6 (57.0-58.2) | 76.2 (75.8-76.6) | 0.66 (0.66-0.67) |
| **5** | 6,595/65,156 | | 18.6 (18.2-19.1) | 95.9 (95.7-96.0) | 70.7 (69.6-71.8) | 68.8 (68.4-69.1) | 0.57 (0.57-0.58) |
| **Centor** | | | | | | | |
| **≥ 1** | 6,647/52 | 41.6 (40.5-42.8) | 99.9 (99.7-100.0) | 1.3 (1.0-1.7) | 41.9 (40.8-43.1) | 96.2 (86.8-99.5) | 0.51 (0.50-0.51) |
| **≥ 2** | 6,375/324 | | 99.0 (98.6-99.4) | 7.6 (6.8-8.5) | 43.3 (42.1-44.6) | 91.7 (88.1-94.4) | 0.53 (0.53-0.54) |
| **≥ 3** | 5,518/1,181 | | 91.7 (90.6-92.7) | 24.3 (22.9-25.7) | 46.4 (45.0-47.7) | 80.4 (78.0-82.6) | 0.58 (0.57-0.59) |
| **4** | 1,258/5,441 | | 28.5 (26.8-30.2) | 88.1 (87.1-89.1) | 63.1 (60.4-65.8) | 63.3 (62.0-64.6) | 0.58 (0.57-0.59) |
| **6-10 years** | | | | | | | |
| **FeverPAIN** | | | | | | | |
| **≥ 1** | 10,835/70 | 44.6 (43.7-45.6) | 99.9 (99.8-100.0) | 1.1 (0.9-1.4) | 44.9 (43.9-45.8) | 95.7 (88.0-99.1) | 0.51 (0.50-0.51) |
| **≥ 2** | 10,481/424 | | 98.6 (98.3-98.9) | 5.9 (5.3-6.5) | 45.8 (44.8-46.7) | 84.2 (80.4-87.5) | 0.52 (0.52-0.53) |
| **≥ 3** | 7,711/3,194 | | 84.3 (83.3-85.3) | 40.2 (39.0-41.5) | 53.2 (52.1-54.3) | 76.1 (74.6-77.6) | 0.62 (0.61-0.63) |
| **≥ 4** | 3,966/6,939 | | 53.5 (52.1-54.9) | 77.4 (76.3-78.5) | 65.6 (64.1-67.1) | 67.4 (66.3-68.5) | 0.65 (0.65-0.66) |
| **5** | 1,120/9,785 | | 18.0 (16.9-19.1) | 96.0 (95.4-96.4) | 78.2 (75.7-80.6) | 59.2 (58.3-60.2) | 0.57 (0.56-0.58) |
| **Centor** | | | | | | | |
| **≥ 1** | 1,037/6 | 56.5 (53.4-59.5) | 99.8 (99.1-100.0) | 1.1 (0.4-2.6) | 56.7 (53.6-59.7) | 83.3 (35.9-99.6) | 0.50 (0.50-0.51) |
| **≥ 2** | 1,004/39 | | 98.8 (97.6-99.5) | 7.0 (4.9-9.8) | 58.0 (54.8-61.0) | 82.1 (66.5-92.5) | 0.53 (0.52-0.54) |
| **≥ 3** | 890/153 | | 91.9 (89.3-93.9) | 23.1 (19.3-27.3) | 60.8 (57.5-64.0) | 68.6 (60.6-75.9) | 0.57 (0.55-0.60) |
| **4** | 280/763 | | 35.8 (31.9-39.8) | 84.8 (81.2-88.0) | 75.4 (69.9-80.3) | 50.5 (46.8-54.1) | 0.60 (0.58-0.63) |
| **11-15 years** | | | | | | | |
| **FeverPAIN** | | | | | | | |
| **≥ 1** | 9,169/84 | 29.2 (28.3-30.1) | 100.0 (99.8-100.0) | 1.3 (1.0-1.6) | 29.4 (28.5-30.4) | 98.8 (93.5-100.0) | 0.51 (0.50-0.51) |
| **≥ 2** | 8,814/439 | | 99.0 (98.5-99.3) | 6.3 (5.7-6.9) | 30.3 (29.4-31.3) | 93.6 (90.9-95.7) | 0.53 (0.52-0.53) |
| **≥ 3** | 5,986/3,267 | | 86.1 (84.7-87.4) | 44.1 (42.9-45.3) | 38.8 (37.6-40.1) | 88.5 (87.3-89.6) | 0.65 (0.64-0.66) |
| **≥ 4** | 2,878/6,375 | | 56.3 (54.4-58.2) | 79.3 (78.3-80.3) | 52.8 (51.0-54.7) | 81.5 (80.5-82.4) | 0.68 (0.67-0.69) |
| **5** | 772/8,481 | | 20.1 (18.6-21.6) | 96.5 (96.0-96.9) | 70.2 (66.8-73.4) | 74.6 (73.6-75.5) | 0.58 (0.57-0.59) |
| **Centor** | | | | | | | |
| **≥ 1** | 769/6 | 35.5 (32.1-39.0) | 100.0 (98.7-100.0) | 1.2 (0.4-2.6) | 35.8 (32.4-39.3) | 100.0 (54.1-100.0) | 0.51 (0.50-0.51) |
| **≥ 2** | 726/49 | | 99.3 (97.4-99.9) | 9.4 (7.0-12.3) | 37.6 (34.1-41.2) | 95.9 (86.0-99.5) | 0.54 (0.53-0.56) |
| **≥ 3** | 619/156 | | 89.8 (85.6-93.1) | 25.6 (21.8-29.7) | 39.9 (36.0-43.9) | 82.1 (75.1-87.7) | 0.58 (0.55-0.60) |

| | | | | | | | |
|---|---|---|---|---|---|---|---|
| **4** | 134/641 | | 28.7 (23.5-34.5) | 89.0 (85.9-91.6) | 59.0 (50.1-67.4) | 69.4 (65.7-73.0) | 0.59 (0.56-0.62) |
| **16-40 years** | | | | | | | |
| **FeverPAIN** | | | | | | | |
| **≥ 1** | 37,909/297 | 36.2 (35.8-36.7) | 99.9 (99.8-99.9) | 1.1 (1.0-1.3) | 36.5 (36.0-37.0) | 94.3 (91.0-96.6) | 0.51 (0.50-0.51) |
| **≥ 2** | 36,613/1,593 | | 98.8 (98.6-99.0) | 5.9 (5.6-6.2) | 37.4 (36.9-37.9) | 89.8 (88.2-91.2) | 0.52 (0.52-0.53) |
| **≥ 3** | 26,529/11,677 | | 86.3 (85.7-86.9) | 40.1 (39.5-40.8) | 45.0 (44.4-45.6) | 83.8 (83.1-84.4) | 0.63 (0.63-0.64) |
| **≥ 4** | 13,832/24,374 | | 56.6 (55.8-57.5) | 75.4 (74.9-76.0) | 56.7 (55.9-57.5) | 75.4 (74.8-75.9) | 0.66 (0.66-0.67) |
| **5** | 4,042/34,164 | | 20.0 (19.3-20.7) | 94.8 (94.5-95.1) | 68.5 (67.1-70.0) | 67.6 (67.1-68.1) | 0.57 (0.57-0.58) |
| **Centor** | | | | | | | |
| **≥ 1** | 3,476/24 | 41.9 (40.3-43.6) | 99.9 (99.6-100.0) | 1.1 (0.7-1.7) | 42.2 (40.5-43.8) | 95.8 (78.9-99.9) | 0.51 (0.50-0.51) |
| **≥ 2** | 3,360/140 | | 99.3 (98.7-99.6) | 6.3 (5.3-7.5) | 43.3 (41.6-45.0) | 92.1 (86.4-96.0) | 0.53 (0.52-0.53) |
| **≥ 3** | 2,932/568 | | 92.8 (91.3-94.0) | 22.7 (20.9-24.6) | 46.4 (44.6-48.2) | 81.3 (77.9-84.5) | 0.58 (0.57-0.59) |
| **4** | 635/2,865 | | 25.6 (23.3-27.9) | 87.2 (85.7-88.6) | 59.1 (55.1-62.9) | 61.9 (60.1-63.7) | 0.56 (0.55-0.58) |
| **≥41 years** | | | | | | | |
| **FeverPAIN** | | | | | | | |
| **≥ 1** | 13,114/273 | 27.0 (26.2-27.7) | 99.5 (99.2-99.7) | 2.6 (2.3-3.0) | 27.4 (26.7-28.2) | 93.8 (90.2-96.3) | 0.51 (0.51-0.51) |
| **≥ 2** | 12,191/1,196 | | 97.1 (96.6-97.7) | 11.2 (10.6-11.8) | 28.8 (28.0-29.6) | 91.4 (89.7-92.9) | 0.54 (0.54-0.55) |
| **≥ 3** | 7,253/6,134 | | 78.3 (76.9-79.7) | 54.7 (53.7-55.7) | 39.0 (37.9-40.1) | 87.2 (86.4-88.1) | 0.67 (0.66-0.67) |
| **≥ 4** | 2,878/10,509 | | 44.4 (42.7-46.0) | 86.9 (86.3-87.6) | 55.7 (53.8-57.5) | 80.9 (80.1-81.6) | 0.66 (0.65-0.67) |
| **5** | 661/12,726 | | 13.2 (12.1-14.3) | 98.1 (97.8-98.4) | 72.0 (68.4-75.4) | 75.4 (74.6-76.1) | 0.56 (0.55-0.56) |
| **Centor** | | | | | | | |
| **≥ 1** | 1,365/16 | 33.2 (30.8-35.8) | 100.0 (99.2-100.0) | 1.7 (1.0-2.8) | 33.6 (31.1-36.2) | 100.0 (79.4-100.0) | 0.51 (0.50-0.51) |
| **≥ 2** | 1,285/96 | | 98.5 (96.9-99.4) | 9.7 (7.8-11.7) | 35.2 (32.6-37.9) | 92.7 (85.6-97.0) | 0.54 (0.53-0.55) |
| **≥ 3** | 1,077/304 | | 89.1 (85.9-91.8) | 27.5 (24.7-30.6) | 38.0 (35.1-41.0) | 83.6 (78.9-87.5) | 0.58 (0.56-0.60) |
| **4** | 209/1,172 | | 28.1 (24.0-32.5) | 91.3 (89.3-93.1) | 61.7 (54.8-68.3) | 71.8 (69.2-74.4) | 0.60 (0.57-0.62) |

Above/Below = consultations above FeverPAIN threshold/consultations below FeverPAIN threshold. PPV = positive predictive value. NPV = negative predictive value. AUROC = area under receiver operating characteristic curve.

**Supplementary Figure 1.** Proportion of treated and untreated patients if FeverPAIN ≥4 alone was used to guide antibiotic supply in 100 patients presenting with sore throat to the STTT service by age group.

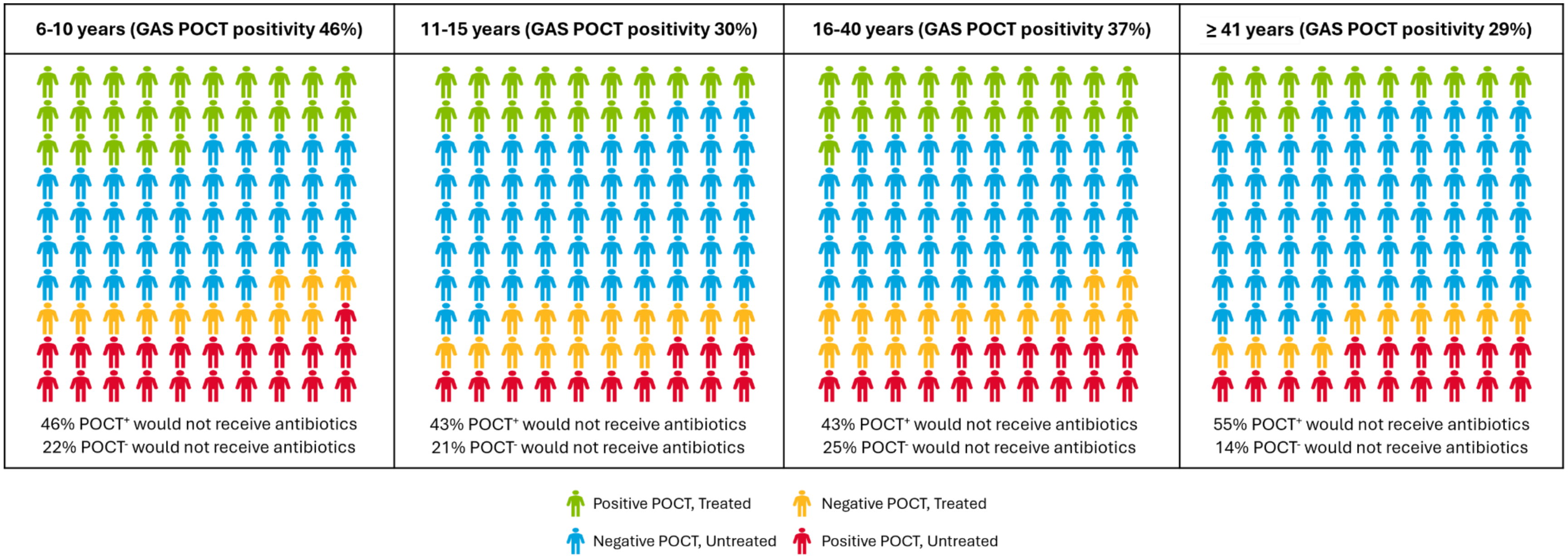


POCT$^{+}$ = POCT-positive. POCT$^{-}$ = POCT-negative.
